# Evolving Epidemiology and Impact of Non-pharmaceutical Interventions on the Outbreak of Coronavirus Disease 2019 in Wuhan, China

**DOI:** 10.1101/2020.03.03.20030593

**Authors:** Chaolong Wang, Li Liu, Xingjie Hao, Huan Guo, Qi Wang, Jiao Huang, Na He, Hongjie Yu, Xihong Lin, An Pan, Sheng Wei, Tangchun Wu

**Affiliations:** Department of Epidemiology and Biostatistics, Ministry of Education Key Laboratory of Environment and Health, and State Key Laboratory of Environmental Health (Incubating), School of Public Health, Tongji Medical College, Huazhong University of Science and Technology, Wuhan, China; Department of Occupational and Environmental Health, Ministry of Education Key Laboratory of Environment and Health, and State Key Laboratory of Environmental Health (Incubating), School of Public Health, Tongji Medical College, Huazhong University of Science and Technology, Wuhan, China; School of Public Health, Ministry of Education Key Laboratory of Public Health Safety, Fudan University, Shanghai, China; Department of Biostatistics, Harvard T.H. Chan School of Public Health, Boston, MA, USA

## Abstract

**BACKGROUND:** We described the epidemiological features of the coronavirus disease 2019 (Covid-19) outbreak, and evaluated the impact of non-pharmaceutical interventions on the epidemic in Wuhan, China.

**METHODS:** Individual-level data on 25,961 laboratory-confirmed Covid-19 cases reported through February 18, 2020 were extracted from the municipal Notifiable Disease Report System. Based on key events and interventions, we divided the epidemic into four periods: before January 11, January 11-22, January 23 - February 1, and February 2-18. We compared epidemiological characteristics across periods and different demographic groups. We developed a susceptible-exposed-infectious-recovered model to study the epidemic and evaluate the impact of interventions.

**RESULTS:** The median age of the cases was 57 years and 50.3% were women. The attack rate peaked in the third period and substantially declined afterwards across geographic regions, sex and age groups, except for children (age <20) whose attack rate continued to increase. Healthcare workers and elderly people had higher attack rates and severity risk increased with age. The effective reproductive number dropped from 3.86 (95% credible interval 3.74 to 3.97) before interventions to 0.32 (0.28 to 0.37) post interventions. The interventions were estimated to prevent 94.5% (93.7 to 95.2%) infections till February 18. We found that at least 59% of infected cases were unascertained in Wuhan, potentially including asymptomatic and mild-symptomatic cases.

**CONCLUSIONS:** Considerable countermeasures have effectively controlled the Covid-19 outbreak in Wuhan. Special efforts are needed to protect vulnerable populations, including healthcare workers, elderly and children. Estimation of unascertained cases has important implications on continuing surveillance and interventions.

## INTRODUCTION

The Coronavirus Disease 2019 (Covid-19) is an emerging respiratory infectious disease caused by SARS-CoV-2 (also known as 2019-nCoV), which first occurred in early December 2019 in Wuhan, China. Until March 1, Covid-19 has affected more than 79,900 individuals and caused 2873 deaths in China, and quickly spread to over 55 countries worldwide.^1^ Although some studies with varying sample sizes have described the clinical characteristics of patients with Covid-19,^2-7^ and a previous study has reported the early transmission dynamics of the first 425 confirmed cases in Wuhan,^8^ most recent data are required to illustrate the full spectrum of the epidemiological characteristics of the outbreak in Wuhan.

During the outbreak, the Chinese authorities have implemented a series of non-pharmaceutical interventions to control the epidemic (details in **Fig. 1**), including an unprecedented policy of *cordon sanitaire* in Wuhan City on January 23, 2020, severely restricting outbound traffic and affecting about 10 million people. Wuhan is a transportation hub in central China with massive human movement before the quarantine policy, especially because of the approaching of the Chinese New Year. Several modelling studies have used the international cases exported from Wuhan to extrapolate the severity of epidemic in Wuhan, which estimated much larger numbers of infected cases than those officially reported, implying a substantial amount of unascertained cases.^9,10^ While the huge discrepancy remained unexplained, these early models can no longer be applied since January 23 due to the intensive intra-city and inter-city traffic restriction, social distancing measures, and improvement of medical resources within Wuhan city. These interventions would inevitably affect the model parameters such as the transmission rate across time. In addition, many previous modeling studies have used the date of laboratory confirmation in the analysis without considering the long lag between onset and confirmation date for the early cases.^11-13^ Moreover, several recent studies have reported a nonnegligible proportion of asymptomatic cases^14-16^ and transmissibility of the asymptomatic or presymptomatic cases,^17-19^ which were not considered by previous models.

**Figure 1.**
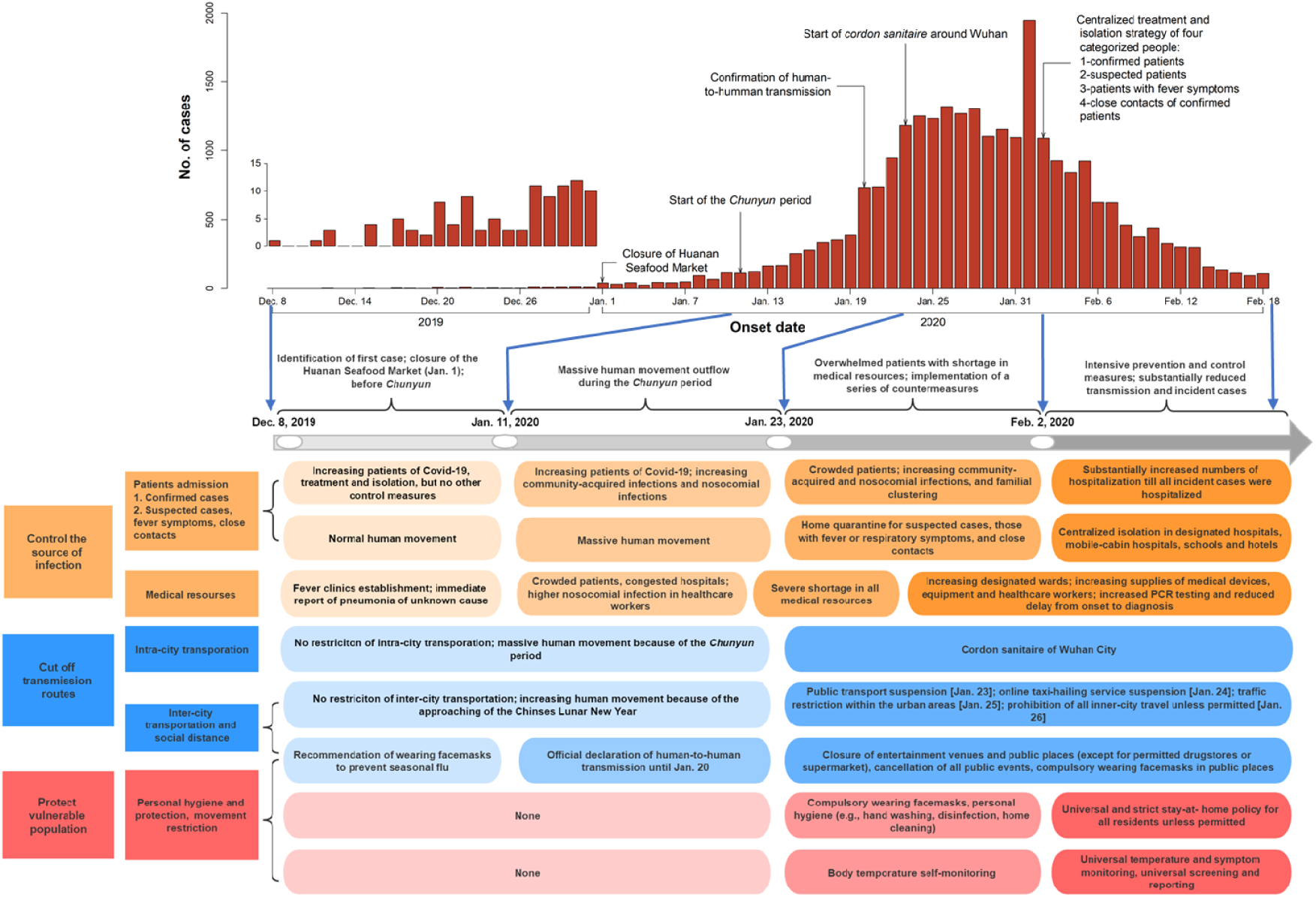
The daily Covid-19 onset and the control measures across different periods.

In this study, we described the epidemiological characteristics of the laboratory-confirmed patients with Covid-19 in Wuhan till February 18, 2020. We developed a novel susceptible-exposed-infectious-recovered (SEIR) model to study the epidemic by accounting for time-varying population movement, ascertainment rate, transmission rate, and duration from illness onset to hospitalization. We compared model prediction under different scenarios and the actual reported cases to evaluate the overall impact of the city quarantine and subsequent interventions on the epidemic in Wuhan.

## METHODS

### Source of data

Covid-19 cases from December 2019 till February 18, 2020 were extracted on February 19 from the municipal Notifiable Disease Report System, including the general information of birth date, sex, occupation, residential district, date of illness onset (the self-reported date of symptoms such as fever, cough, or other respiratory symptoms), and date of confirmed diagnosis (the laboratory confirmation date of SARS-CoV-2 in the bio-samples). The identifiable personal information was removed for privacy protection. A case was recorded as a healthcare worker if reported to work in a hospital or clinic.

### Case definitions

Cases were diagnosed and the severity status was categorized as mild, moderate, severe, and critical according to the Diagnosis and Treatment Scheme for Covid-19 released by the National Health Commission of China.^20^ A laboratory-confirmed case was defined if the patient had a positive test of SARS-CoV-2 virus by the real-time reverse-transcription-polymerase-chain-reaction (RT-PCR) assay or high-throughput sequencing of nasal and pharyngeal swab specimens. We only included laboratory-confirmed cases in our analyses for consistency of case definition throughout the periods.

### Classification of four time periods

To better reflect the dynamics of the Covid-19 epidemic and corresponding interventions, we classified the outbreak into four periods based on important dates that could affect the virus transmission (**Fig. 1**). The time before January 11, 2020, the first date of *Chunyun* (massive migration for the Chinese New Year), was considered as the first period when no intervention was imposed. The second period referred to the *Chunyun* of January 11-22, 2020, when massive population movement occurred and was expected to accelerate the spread of Covid-19. No strong intervention was imposed except for the announcement of human-to-human transmission and infections in healthcare workers on January 20. The hospitals started to be overcrowded with people with fever or respiratory symptoms. During the third period between January 23 and February 1, the local government first blocked all outbound transportations on January 23 and subsequently suspended public transit and banned all vehicular traffic within the city. Other social distancing measures were also implemented, including compulsory mask-wearing in public places and cancellation of social gathering. Due to severe shortage of medical resources in this period, many confirmed or suspected cases could not receive timely treatment and were self-quarantined at home. On February 2, with improvement in medical resources, the government implemented the policy of centralized quarantine and treatment of all confirmed and suspected cases, those with fever or respiratory symptoms, as well as close contacts of confirmed cases in designated hospitals or facilities. Meanwhile, temperature monitoring and stay-at-home policies were implemented to all residents in the city. Taken together, we divided the outbreak in Wuhan into four periods (before January 11, January 11-22, January 23-February 1, and February 2-18, 2020, respectively) with specific intervention activities provided in **Fig. 1**.

### Statistical analysis

We calculated the daily onset numbers of confirmed Covid-19 cases from December 2019 till February 18, 2020. We estimated the attack rate, defined as the number of infections per day per 10^6^ people, by age, sex, healthcare occupation, and residential districts, with the subtotal population size in each stratum from the Wuhan Statistical Yearbook 2018. Logistic regression was used to evaluate the association of age, sex, time period and healthcare occupation with the disease severity (mild/moderate versus severe/critical). Odds ratios (ORs) were reported along with the 95% confidence intervals (CIs) and *P* values.

We extended the classic SEIR model to account for population movement, unascertained cases, and quarantine by hospitalization (**Fig. 2** and **Supplementary Methods**). We chose to analyze data from January 1, 2020, when the Huanan Seafood Market was closed. We assumed a constant population size of 10 million of Wuhan with equal daily inbound and outbound travelers (500,000 for January 1-10, 800,000 for January 11-22 due to *Chunyun*, and 0 afterwards due to *cordon sanitaire* since January 23).^9^ We divided the population into six compartments including susceptible individuals, latent cases, ascertained cases, unascertained cases, hospitalized cases, and removed individuals. Here, unascertained cases included asymptomatic cases and those with mild symptoms who could recover without seeking medical care and thus were not reported to authorities. We assumed only those seeking medical care would be reported and quarantined by hospitalization. Dynamics of these six compartments across time were described by ordinary differential equations (**Supplementary Methods)**, along with the key parameters. The daily case data were assumed to the Poisson regression under the SEIR model. Considering the impacts of major interventions, we assumed that the transmission rate and ascertainment rate were the same in the period 1 and 2, while the two parameters were different for the period 3 and 4. The effective reproductive number *R*_*t*_, defined as the expected number of secondary cases infected by a primary case, was computed for each period.

**Figure 2.**
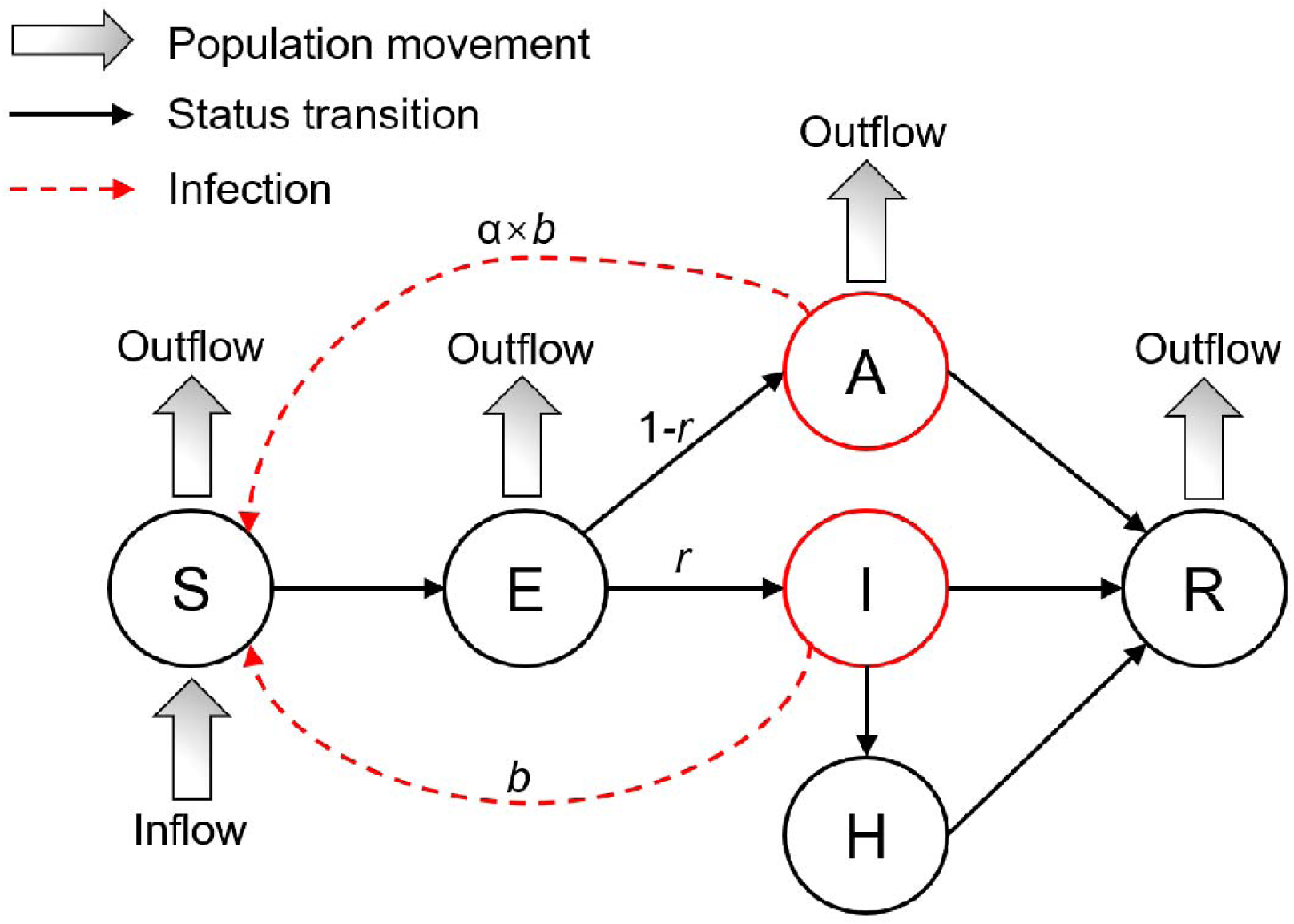
Illustration of the extended susceptible-exposed-infectious-recovered model. We divided the population in Wuhan into six compartments: S (susceptible), E (latent), I (reported infections), A (unreported infections), H (hospitalized), and R (removed). Two key parameters in the model are *r* (ascertainment rate) and *b* (transmission rate). The details of the model assumptions and dynamics of these six compartments across time are described in the Supplementary Methods.

Initial states of the SEIR model and parameter settings for the main and sensitivity analyses are described in detail in the **Supplementary Methods**. The transmission rate and ascertainment rate in different periods and their 95% Credible Intervals (CrIs) were estimated by fitting the observed data from January 1 to February 10 using Markov Chain Monte Carlo (MCMC, **Supplementary Methods**).We used the fitted model to predict the trend from February 11 to 18 and compared that with the observed data to assess the accuracy of the model.

### Ethics Approval

The ethics approval was considered exempt because all date collection and analyses belong to a part of continuing public health outbreak investigation determined by the National Health Commission of China.

## RESULTS

### General characteristics of patients with Covid-19

Our analyses included a total of 25,961 confirmed cases, among whom 49.7% were men and 50.3% were women (**Table 1**). The epidemic curve according to the onset date and key interventions is shown in the **Fig. 1**. Most cases occurred between January 20 and February 6, with a spike on February 1. There was a substantial delay between the onset date and confirmation date in early periods, with the lag decreasing over periods (median 22, 14, 10 and 5 days for the four periods, respectively; **Fig. S1**).

**Table 1.** The numbers and proportions (%) of the laboratory-confirmed Covid-19 cases stratified by sex and age in Wuhan from December 8, 2019 to February 18, 2020

| Characteristics | Before January 11 | January 11-22 | January 23 - February 1 | February 2-18 | Total |
| --- | --- | --- | --- | --- | --- |
| **Total no.** | 637 | 4599 | 12879 | 7846 | 25961 |
| **Sex — no. (%)** |  |  |  |  |  |
| **Male** | 334 (52.4) | 2266 (49.3) | 6354 (49.3) | 3952 (50.4) | 12906 (49.7) |
| **Female** | 303 (47.6) | 2333 (50.7) | 6525 (50.7) | 3894 (49.6) | 13055 (50.3) |
| **Median age (IQR) - yr** | 60.9 (19.4) | 57.1 (21.3) | 57.2 (22.2) | 56.4 (24.5) | 57.0 (22.7) |
| **Age group — no. (%)** |  |  |  |  |  |
| **0-19 yr** | 2 (0.3) | 16 (0.3) | 79 (0.6) | 193 (2.5) | 290 (1.1) |
| **20-39 yr** | 71 (11.1) | 820 (17.8) | 2235 (17.4) | 1508 (19.2) | 4634 (17.8) |
| **40-59 yr** | 235 (36.9) | 1742 (37.9) | 4929 (38.3) | 2851 (36.3) | 9757 (37.6) |
| **60-79 yr** | 287 (45.1) | 1806 (39.3) | 5026 (39.0) | 2826 (36.0) | 9945 (38.3) |
| **≥80 yr** | 42 (6.6) | 215 (4.7) | 610 (4.7) | 468 (6.0) | 1335 (5.1) |
| **Healthcare workers — no. (%)** | 26 (4.1) | 411 (8.9) | 632 (4.9) | 247 (3.1) | 1316 (5.1) |

The outbreak started from the urban districts and gradually spread to the suburban and rural areas across the four periods, leading to strong geographic differences with the highest attack rates in the urban districts (**Fig. S2**). The average daily attack rate per 10^6^ people dramatically increased from 2.2 (95% CI, 2.0 to 2.4) before January 11, to 44.9 (43.6 to 46.2) between January 11 and 22, and to 150.9 (148.3 to 153.5) between January 23 and February 1, while dropped to 54.1 (52.9 to 55.3) after February 2 (**Fig. 3A**). Similar patterns were observed for men and women, with slightly higher attack rate in women (**Fig. 3A**). A total of 1316 healthcare workers were infected, representing 5.1% of the total cases (**Table 1**). The average attack rate in local healthcare workers (144.7 per 10^6^ people; 95% CI, 137.0 to 152.8) was substantially higher than that in the general population (41.7 per 10^6^ people; 41.2 to 42.2) overall, and particularly in the third period (507.4 per 10^6^ people; 468.6 to 548.5; **Fig. 3A**).

**Figure 3.**
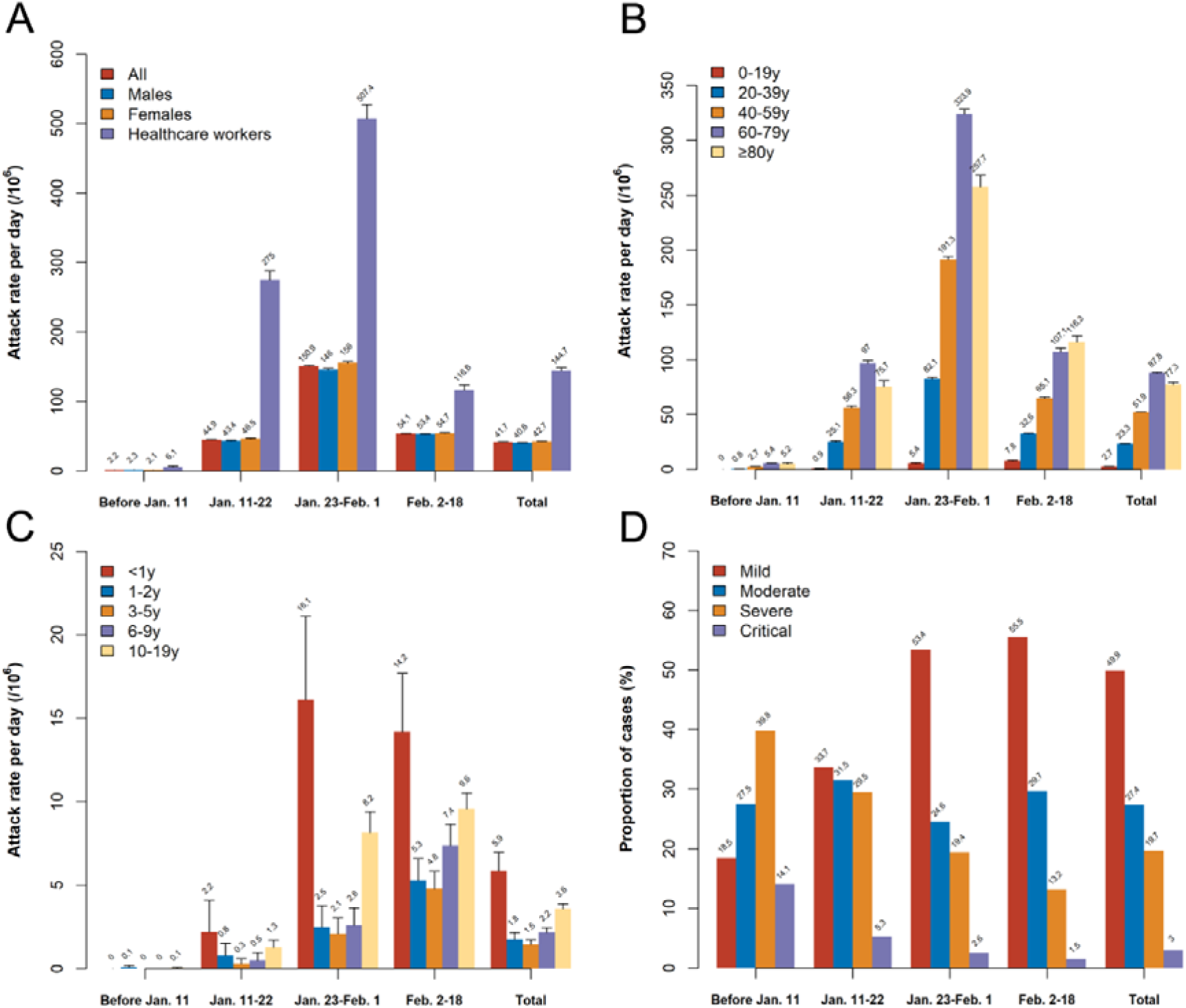
Attack rates in different groups and proportion of severity groups across the four periods. (A) Attack rates in all patients, men, women and healthcare workers; (B) Attack rates in different age groups; (C) Attack rates in age groups younger than 20 years; (D) Proportion of severity groups.

The median age of the patients was 57 years, with the majority (n=19,693, 75.9%) aged 40 to 79 years (**Table 1**). The attack rate peaked in the third period then declined in the fourth period for those older than 20 years, while it continued to increase throughout the periods for children (age <20 years) (**Fig. 3B**), particularly for the infants below 1 year old (**Fig. 3C**).

The clinical severities of the confirmed cases (n=25,727) were classified into mild (n=12,834, 49.9%), moderate (n=7052, 27.4%), severe (n=5071, 19.7%) and critical (n=770, 3.0%; **Fig. 3D**). The proportion of severe/critical cases decreased gradually over time, accounting for 53.9%, 34.8%, 22.0% and 14.7% of the classifiable cases in the four periods, respectively, while the proportion of mild cases increased dramatically (**Fig. 3D**). Compared to cases aged 20 to 40, children younger than 10 were less likely to be severe/critical (OR, 0.33; 95% CI, 0.14 to 0.82), while the ORs increased with age: 1.44 (1.30 to 1.60), 2.76 (2.49 to 3.05), and 5.11 (4.42 to 5.91) for cases aged 40-60, 60-80, or ≥80, respectively (**Table S1**). In addition, females were at lower risk of severity than males (OR, 0.89; 0.84 to 0.95), while there was weak evidence that healthcare workers were at higher risk of severity (OR, 1.12; 0.96 to 1.31).

### Modeling the epidemic trend in Wuhan

Our SEIR model fit the observed data well, except for the outlier on February 1 (**Fig. 4A**). The slight overprediction for the last five days (February 14-18) was likely due to the delay in laboratory confirmation of recent cases. The transmission rate decreased from 1.75 (95% CrI, 1.71 to 1.80) before January 23 to 0.58 (0.56 to 0.60) and 0.15 (0.13 to 0.17) after January 23 and February 2, respectively (**Table S2**), which could be translated into *R*_*t*_ of 3.88 (3.77 to 3.99), 3.86 (3.74 to 3.97), 1.26 (1.21 to 1.31), and 0.32 (0.28 to 0.37) for the four periods, respectively (**Fig. 4B**). We estimated the number of cumulative ascertained cases till February 18 to be 49,943 (95% CrI, 43,577 to 56,635) if the trend of the third period was assumed (**Fig. 4C**), or 474,897 (410,660 to 537,448) if the trend of the second period was assumed (**Fig. 4D**), both were much higher than the current ascertained case number of 25,961. These numbers were translated to a total of 48.0% (40.7 to 54.3%) and 94.5% (93.7-95.2%) prevented cases by the interventions.

**Figure 4.**
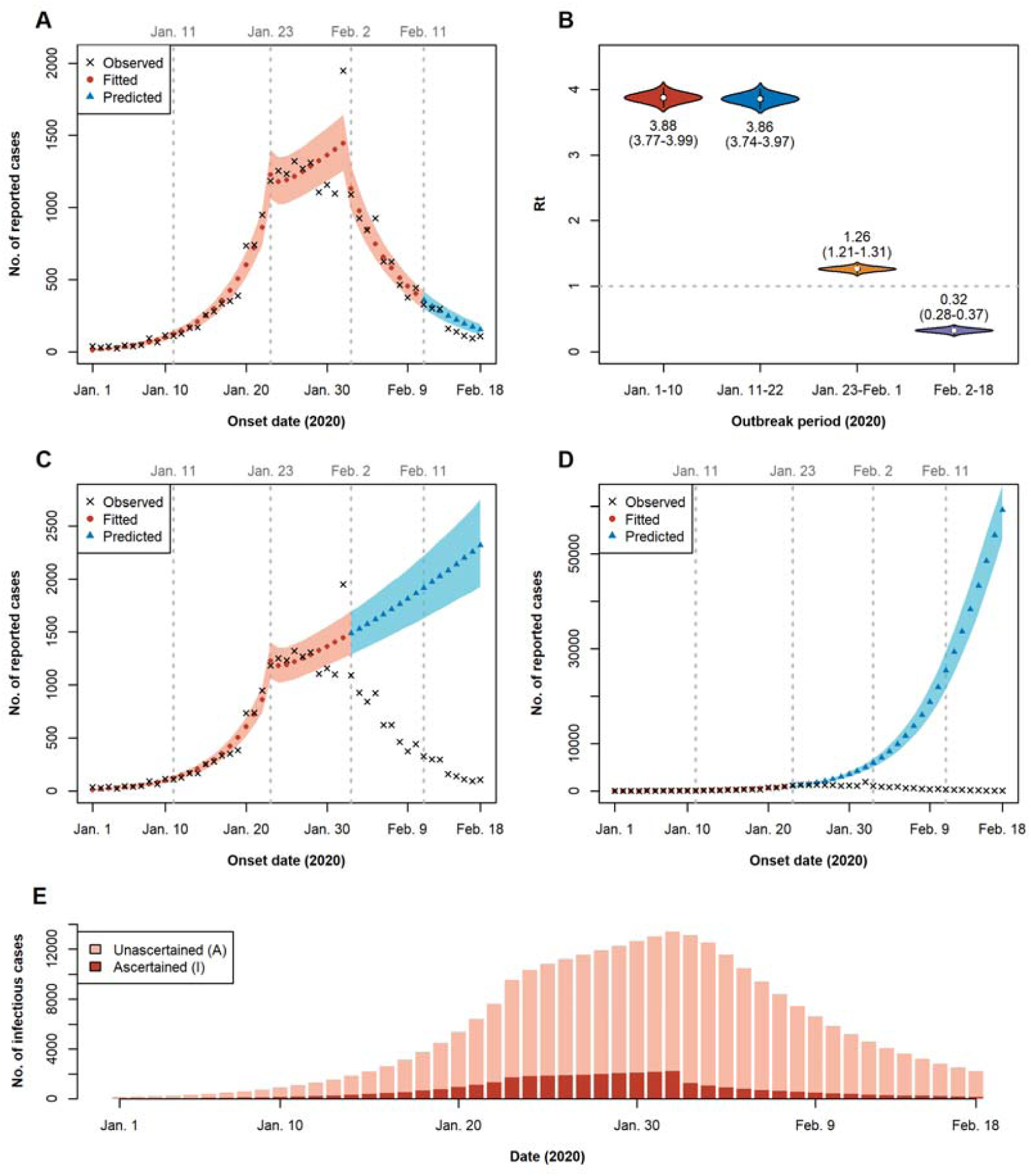
Four-period susceptible-exposed-infectious-recovered modeling of the Covid-19 epidemics in Wuhan. Parameters were fitted based on data from January 1 to February 10. (A) Prediction of cases in February 11-18 (blue) using parameters from February 2-10 (red). (B) Estimated *R*_*t*_ for the four periods (January 1-10, January 11-22, January 23 - February 1, and February 2-18). The mean and 95% credible interval (in parentheses) are labeled below or above the violin plots. (C) Prediction of the fourth period (February 2-18) using parameters from the third period (January 23 - February 1). (D) Prediction of the fourth period (February 2-18) using parameters from the second period (January 11-20). The shaded areas in (A, C and D) are 95% credible intervals of the fitted/predicted values. (E) Estimated number of active infectious cases in Wuhan from January 1 to February 18.

Strikingly, we estimated that the overall ascertainment rate was 0.21 (95% CrI, 0.18-0.24), and similar across the periods (**Table S2**). We predicted the cumulative number of ascertained cases to be 26,252 (95% CrI, 23,116 to 29,522) by February 18, close to the actual reported number of 25,961, while the estimated cumulative number of total cases was 125,959 (105,060 to 151,612). Our model suggested the number of active infectious cases in Wuhan peaked on February 1, and then gradually dropped afterwards (**Fig. 4E**). If the trend remained unchanged, we predicted the number of ascertained cases to become zero by April 22 (95% CrI, April 5 to May 19), 2020, and the total number of both ascertained and unascertained cases would become zero around May 4 (April 17 to May 30), 2020.

We performed a series of sensitivity analyses to test the robustness of our results by considering the outlier data point on February 1, and varying incubation and infectious periods, transmission ratio between unascertained and ascertained cases, and initial values of the model (**Fig. S4-S12, Tables S2-S4**). Our major findings of remarkable decrease in *R*_*t*_ in response to interventions and the existence of a large proportion of unascertained cases remained in all sensitivity analyses. In particular, we noticed that the estimated ascertainment rates increased with decreasing initial number of unascertained cases (**Fig. S9-S11, Table S2**). If we assumed an extreme scenario with no unascertained cases initially (**Fig. S11**), the overall ascertainment rate would be 0.41 (0.36-0.47), which would be the upper bound of the ascertainment rate. We also tested a simplified model assuming no unascertained cases anytime, but this simplified model performed significantly worse than the full model in fitting the data (**Fig. S12**).

## DISCUSSION

Here we provide a comprehensive assessment of the epidemiological characteristics of the laboratory-confirmed Covid-19 cases in Wuhan, the epicenter of outbreak. The virus affected equally to men and women and most cases were middle-aged and elderly adults.

The attack rate continued to increase before February 2 while dramatically declined thereafter for all groups, except for children (age <20 years). Consistent with early analyses, younger people were less likely to be affected,^8,11,21^ but we found that the attack rate continued to increase over time for those aged under 20 years. In particularly, infants under the age of 1 year had the highest attack rate than the other age groups of children, probably because they cannot wear masks and have low immunity.^22^ Children had a lower chance of getting infected probably because they had less frequent social activities during the school winter break starting in early or middle January, but the attack rate increased when all people were required to stay at home and risk of familial clustering of infection started to increase.^11^ Our results also indicated that healthcare workers and elderly people had higher attack rates and the severity increased significantly with age. Therefore, special attention and efforts should be applied to protect and reduce transmission and progression in vulnerable populations including healthcare workers, elderly people and children.

Despite that the outbreak started in early December, no strong interventions were taken before January 20 when the human-to-human transmission was officially announced. The outbreak quickly spread from the urban areas to the suburban and rural areas. The attack rate in the healthcare workers was substantially higher between January 11 and February 1, indicating a high risk of nosocomial infections. This was probably due to lower awareness of protection before January 20, and later severe shortage of medical resources including designated wards and personal protective equipment in hospitals confronting overwhelmed patients.

We compared our model prediction with published modeling studies using independent datasets. Based on early international exported cases, Wu et al.^9^ estimated that 75,815 (95% CrI, 37,304 to 130,330) individuals had been infected in Greater Wuhan as of January 25, 2020. For comparison, we estimated the number to be 36,798 (95% CrI, 30,898 to 43,390) by the same day, including both ascertained and unascertained cases. The discrepancy was mainly due to different assumptions of population size, which was 19 million for the Greater Wuhan Area including surrounding cities by Wu et al.^9^ versus 10 million for the Wuhan city in our analysis. After accounting for the population size, the estimates of prevalence were indeed highly consistent (0.40% versus 0.37%). Another study,^10^ which was also based on international exported cases but used a different model with an assumed population size of 11 million, estimated the number of infected cases in Wuhan to be 29,500 (14,300 to 85,700) on January 23, 2020, closely matching our prediction of 26,144 cases (21,936 to 30,748) on the same day. These studies supported the validity of key assumptions made in our main analysis, including the initial ascertainment rate of 0.5. For example, if we assumed no unascertained cases in the initial state (**Fig. S10**), our estimated cumulative number of cases would be 13,441 (10,853 to 16,356) on January 23 and 18,826 (15,115 to 23,039) on January 25, much lower than those estimated by these two studies.

Our finding of substantial unascertained cases has important implications for the evaluation and control of the Covid-19 epidemic.^23^ These unascertained cases were likely asymptomatic or with mild symptoms, who could mostly recover without seeking medical care. There is accumulating evidence on the existence of many asymptomatic or presymptomatic cases. For example, asymptomatic cases were estimated to account for 34.6% of the virus positive cases onboard the Princess Cruise ship.^14^ Several recent reports also highlighted the difficulty to detect Covid-19 cases: about two thirds of the cases exported from mainland China remained undetected worldwide,^24^ and the detection capacity varied from 11% in low surveillance countries to 40% in high surveillance countries.^25,26^ Consistent with these studies, our analyses and extensive simulations also indicated an ascertainment rate of 14%-41% in Wuhan (**Table S2**). Increasing evidence also suggested that the asymptomatic and presymptomatic cases can be infectious to the susceptible population,^17-19^ imposing a substantial challenge to the epidemic control.^27^ There would be about two weeks delay in the predicted ending date of the epidemic when taking the unascertained cases into account (**Table S4**). Therefore, understanding the proportion of unascertained cases and the rate of asymptomatic spread will be critical for pandemic prevention of Covid-19, including prioritization the surveillance and control measures.^23,28^

We demonstrated that the series of interventions has been highly effective in controlling the epidemic in Wuhan. Our estimate of *R*_*t*_=3.88 for the first period reflected the basic reproductive number *R*_*0*_ as few interventions had been implemented by then. Some previous studies have reported varied *R*_*0*_ (range 1.40 to 6.49 with a mean of 3.28) due to different data sources, time periods and statistical methods.^29^ Even using the same dataset of the first 425 patients in Wuhan, an early study reported a *R*_*0*_ of 2.20 based on the growth rate of the epidemic curve and the serial interval,^8^ while a recent analysis based on a transmission network model reported a *R*_*0*_ of 3.58, similar to our estimate. The transmissibility was higher than that for the SARS-CoV in 2003 (from 2.2 to 3.6),^30^ and was consistent with the rapid spreading of Covid-19. Nevertheless, by taking drastic social distancing measures and policies of controlling the source of infection, with the tremendous joint efforts from the government, healthcare workers, and the people (**Fig. 1**), *R*_*t*_ was substantially reduced to 0.32 in Wuhan after February 2, which was encouraging for the global efforts fighting against the Covid-19 outbreak using traditional non-pharmaceutical measures.^31^

Some limitations of this study need to be noted. First, while our model prediction aligned well with the observed data, we set the values of several parameters based on earlier epidemiological studies without accounting for the uncertainty,^8,9^ which might reduce the accuracy of our results. Second, we need field investigations and serologic studies to confirm our estimate of the ascertainment rate, and the generalizability to other places is unknown. This may depend on the detection capacity in different locations.^26^ Third, due to the delay in laboratory tests, we might have missed some cases and therefore underestimated the ascertainment rate, especially for the last period. Finally, the impact of the interventions should be considered as a whole and we could not evaluate individual strategies by the epidemic curve.

Taken together, both the epidemiological characteristics and our modeling estimates demonstrated that the aggressive disease containment efforts, including isolation of the source of infection, contact tracing and quarantine, social distancing, and personal protection and prevention, have considerably changed the course of Covid-19 outbreak in Wuhan, when there was neither effective drug nor vaccine for this new infectious disease with high transmission. Our analyses of different periods also have important implications for other countries, where there is a sharp surge in Covid-19 cases and at the early stage of the epidemic,^1^ to combat the outbreak. With ready preparedness, prompt response and evidence-based strategies, the global community can be united to battle the seemly unstoppable pandemic.

## Data Availability

Not available

## Acknowledgement

We are grateful for all staff at the national, provincial and municipal Center for Disease Control and Prevention for providing the data and all medical staff members and field workers who are working on the front line of caring for the patients and collecting the data. We also thank the government at all levels and all citizens in Wuhan for their sacrifice and enormous efforts in battling with the Covid-19.

## Declaration of interests

We declare no competing interests.

## Author contributions

CW, LL, XH and HG contributed equally to the study and are joint first authors. AP, SW, and TW are joint senior authors.

CW, AP, SW and TW designed this study. SW and TW were involved in data collection. All authors provided statistical expertise. AP and CW wrote the first draft of the manuscript. All authors contributed to the interpretation of the results and critical revision of the manuscript for important intellectual content and approved the final version of the manuscript. SW and TW had full access to all the data in the study and take responsibility for the integrity of the data and the accuracy of the data analysis.

## Funding sources

This study was partly supported by the Fundamental Research Funds for the Central Universities (2019kfyXMBZ015), the 111 Project (C.W., L.L., X.H., H.G., Q.W., J.H., A.P., S.W., T.W.). X.L. is supported by Harvard University. No other relationships or activities that could appear to have influenced the submitted work. The sponsors have no role in the study design; the collection, analysis, or interpretation of data; the writing of the report; or in the decision to submit the article for publication.

## Notes

### Competing Interest Statement

The authors have declared no competing interest.

